# Childhood bullying as a cumulative health risk: A dose-response analysis of peer victimization and adult mental and behavioral health outcomes in Saudi Arabia

**DOI:** 10.64898/2026.08.17.26360648

**Authors:** Dalal A. Bin Hamdan

**Author notes:** Corresponding author (DAB).

## Abstract

Childhood peer victimization is increasingly recognized as an adverse childhood experience with long-term consequences for population health. Most existing research treats bullying as a binary exposure, obscuring the dose-response mechanisms through which cumulative victimization generates escalating health risks. This cross-sectional secondary analysis examines whether childhood bullying frequency follows a dose-response gradient associated with adverse adult mental and behavioral health outcomes in Saudi Arabia, and whether socioeconomic disadvantage and family context moderate these associations. We used nationally representative Adverse Childhood Experiences International Questionnaire (ACE-IQ) data from Saudi Arabia (full sample: N = 10,156; analytical sample: N = 4,632). Binary logistic regression models adjusted for socioeconomic status, gender, age cohort, parental supervision, and family structure were estimated for five adult health outcomes: physician-diagnosed anxiety, suicidal ideation, sleep disturbance, tobacco smoking, and substance use. A consistent dose-response gradient was observed. Frequent victims showed higher adjusted odds of tobacco smoking (OR = 6.55, 95% CI 5.81–7.32) and substance use (OR = 2.71, 95% CI 2.26–3.31) compared to those never bullied. Religion-targeted verbal victimization was the strongest predictor of suicidal ideation (OR = 3.01) and substance use (OR = 3.24), independent of bullying frequency. Associations were amplified among socioeconomically disadvantaged respondents. Parental supervision was protective against substance use (OR = 0.45) but paradoxically associated with suicidal ideation, interpreted as a reactive parenting effect. These findings establish childhood bullying as a graded cumulative public health risk amplified by structural disadvantage. Prevention strategies must extend beyond school programs to address structural inequalities and integrate family-based and community-level protective factors.

## Introduction

Adverse childhood experiences (ACEs) are among the most well-documented social determinants of population health, with cumulative exposure linked to elevated risks of mental illness, substance use, chronic disease, and premature mortality across the life course [1,2]. Within the ACE framework, childhood peer victimization — encompassing physical aggression, verbal harassment, social exclusion, and online abuse — constitutes a pervasive form of early adversity affecting an estimated one in three school-age children worldwide [3,4]. Despite robust evidence linking bullying to long-term mental and behavioral health outcomes including anxiety, suicidal ideation, sleep disturbance, and substance use [5,6,7], childhood bullying has received comparatively limited attention as a population-level public health priority.

A critical methodological limitation constrains the existing evidence base: the predominant analytical approach treats bullying as a binary exposure — bullied versus not bullied — obscuring the dose-response mechanisms through which cumulative peer victimization may generate escalating health risks [8,9]. If bullying operates as a graded health-risk stressor consistent with models in which chronic social adversity dysregulates hypothalamic-pituitary-adrenal (HPA) axis function and contributes to allostatic load [10], then binary models systematically underestimate the public health burden of repeated victimization.

The theoretical foundation of this study integrates three frameworks. Cumulative disadvantage theory posits that early-life adversities initiate risk amplification processes that widen health disparities over time [11,12]. Life course theory provides a framework for understanding how early adversity becomes embedded in developmental trajectories [13,14,15]. The social determinants of health framework positions bullying within broader systems of social stratification, emphasizing that health inequalities are produced through differential access to material resources and protective environments [16,17,18].

Evidence from the Middle East and Gulf Cooperation Council (GCC) region remains particularly sparse. Saudi Arabia represents an important and underrepresented context: national data document significant associations between ACEs and adult health risks [19,20,21], yet population-level dose-response analyses of bullying as a cumulative health exposure are absent from the Saudi public health literature [20,22].

This study tests three pre-specified hypotheses using the WHO ACE-IQ [2] on a nationally representative Saudi sample: (H1) any childhood bullying exposure is positively associated with higher odds of adverse adult health outcomes compared to no exposure; (H2) increasing bullying frequency is associated with progressively higher odds of adverse outcomes; and (H3) these associations are stronger among socioeconomically disadvantaged respondents and partially attenuated among those with higher parental attention in childhood. Five outcomes are examined: physician-diagnosed anxiety, suicidal ideation, sleep disturbance, tobacco smoking, and substance use.

## Materials and methods

This study followed the STROBE (Strengthening the Reporting of Observational Studies in Epidemiology) reporting guidelines for cross-sectional studies [23] (S1 Checklist).

### Study design

We conducted a retrospective cross-sectional secondary analysis of nationally representative ACE-IQ data from Saudi Arabia.

### Setting

Data were collected in Saudi Arabia in 2013 by the King Abdullah International Medical Research Center (KAIMRC) in collaboration with the National Family Safety Program (NFSP), Ministry of National Guard Health Affairs. Participants were recruited from community settings across multiple regions of Saudi Arabia using a nationally representative probability sampling strategy.

### Participants

The source population comprised adults residing in Saudi Arabia aged 18–60 years at the time of survey administration. Inclusion criteria: adults aged 18–60 who completed the full ACE-IQ survey. No additional exclusion criteria were applied. The total recruited sample was N = 10,156. The analytical subsample comprised N = 4,632 adults reporting any childhood bullying exposure (’few times’ or ’many times’). The 5,524 respondents reporting no bullying were retained as a descriptive reference group. Fig 1 presents the participant flow diagram.

**Fig 1.**
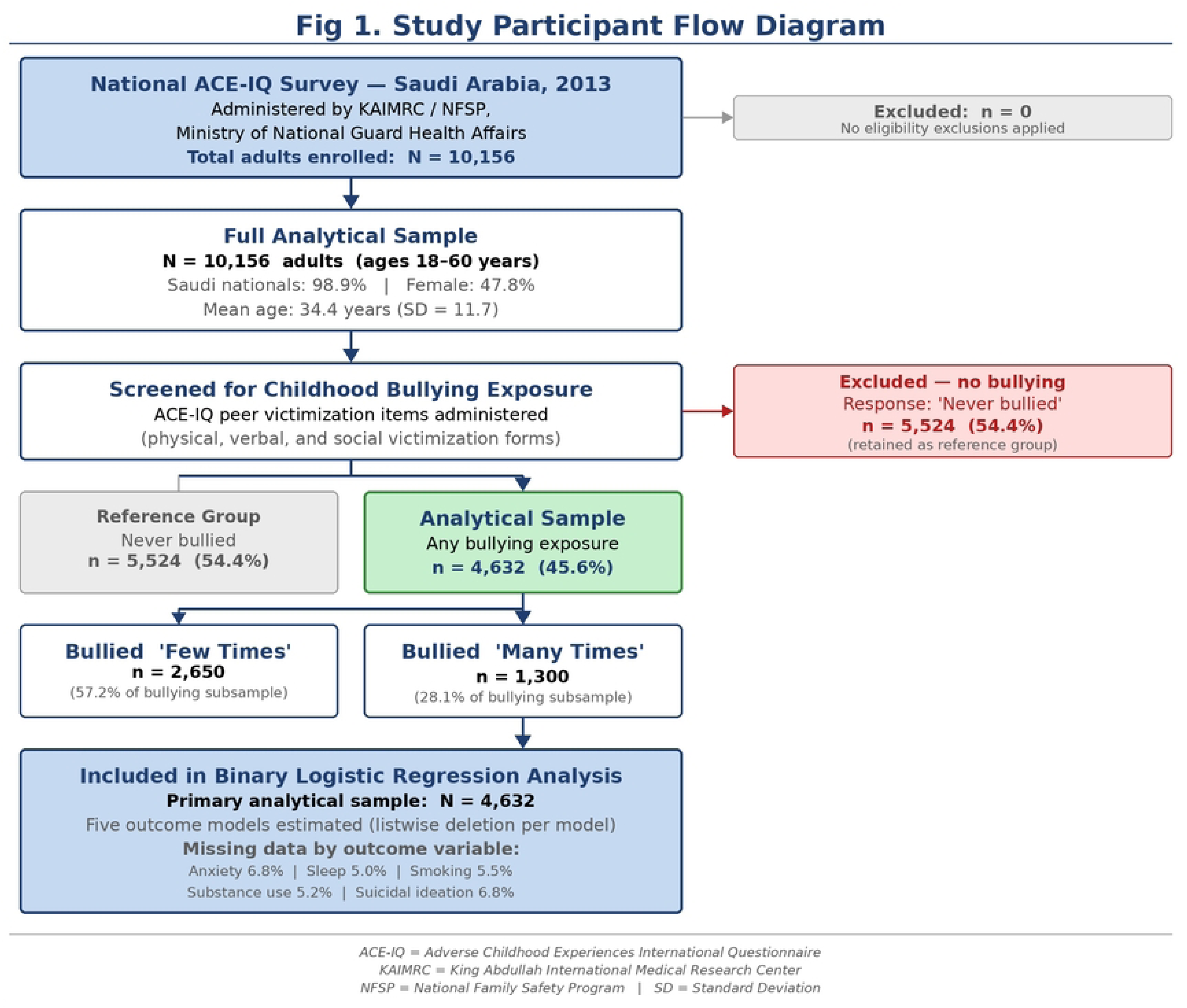
Study participant flow diagram (STROBE Item 13). The national ACE-IQ survey enrolled N = 10,156 adults. Following screening, 5,524 (54.4%) reported never being bullied (reference group) and 4,632 (45.6%) reported any childhood peer victimization (analytical sample), of whom 2,650 (57.2%) reported ’few times’ and 1,300 (28.1%) ’many times’ exposure. ACE-IQ, Adverse Childhood Experiences International Questionnaire; KAIMRC, King Abdullah International Medical Research Center; NFSP, National Family Safety Program.

### Variables

#### Primary exposure: childhood bullying frequency

Childhood bullying exposure was assessed using the peer-violence items of the ACE-IQ [2], capturing: (1) physical victimization (hit, kicked, pushed); (2) verbal victimization (religion-targeted, race/nationality-targeted, appearance-based, sexual harassment); and (3) social victimization (exclusion, relational aggression). Exposure was operationalized as a binary indicator (any vs. none; H1) and as an ordinal frequency variable (few times vs. many times; H2). Reference category in all regression models: ’many times’ bullied.

#### Outcome variables

Five adult health outcomes were analyzed as dichotomous dependent variables: (1) physician-diagnosed anxiety disorder; (2) suicidal ideation (past 6 months); (3) sleep disturbance (past 6 months); (4) tobacco smoking; and (5) substance use.

#### Covariates and effect modifiers

Socioeconomic status was operationalized through educational attainment (less than high school, high school, college or higher), employment status, and marital status. Gender (male/female; male = reference), age cohort (18–31, 32–45 reference, 46–60 years), parental attention (ACE-IQ behavioral monitoring item; 1 = yes, 0 = no), and parental separation or divorce (binary) were included as covariates.

### Data sources and measurement

All variables were drawn from respondents’ self-completed ACE-IQ responses [2]. The ACE-IQ is a validated WHO instrument for cross-national retrospective surveillance of adverse childhood experiences, with established construct validity across multiple cultural settings.

### Bias

Three primary bias sources were addressed. Retrospective recall bias was minimized by the ACE-IQ’s behaviorally specific question wording; residual bias attributable to current health status cannot be excluded and may inflate associations (positive bias direction). Social desirability responding on sensitive items may cause underreporting, producing conservative estimates. Selection bias from the analytical restriction to the bullying subsample is addressed in Table 1.

**Table 1.**
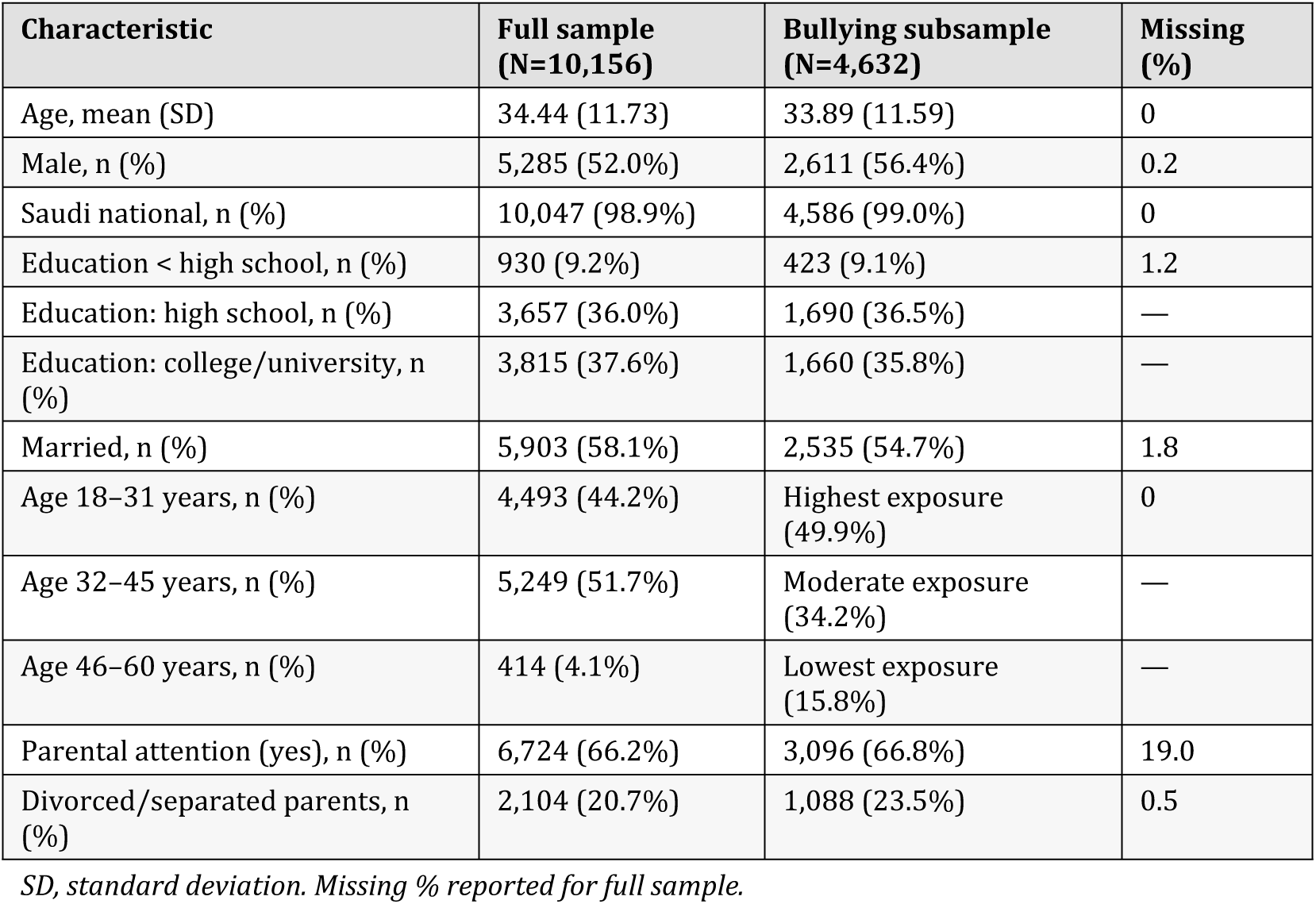
Sample characteristics: full ACE-IQ sample (N = 10,156) and bullying subsample (N = 4,632).

### Study size

The study used a pre-existing nationally representative dataset; no a priori sample size calculation was performed. The analytical sample (N = 4,632) substantially exceeds the 10-events-per-predictor criterion for all models [24].

### Quantitative variables

Age was divided into three life course cohorts (18–31, 32–45, 46–60 years) [12,13]. Educational attainment was grouped into three SES gradient levels. Bullying frequency was retained as an ordinal three-level variable (never, few times, many times) to enable dose-response modeling.

### Statistical analysis

Binary logistic regression models were estimated separately for each of the five adult health outcomes using IBM SPSS Statistics (version 25.0; IBM Corp., Armonk, NY) [24]. Unadjusted (Model 1) and adjusted (Model 2) analyses were conducted. Dose-response patterns (H2) were examined with bullying frequency as an ordinal predictor (’many times’ as reference category). Interaction terms between bullying frequency and socioeconomic status were specified to test H3. Results are reported as adjusted odds ratios (ORs) with 95% confidence intervals (CI). Statistical significance was set at *α* = 0.05 (two-tailed). Multicollinearity was assessed using variance inflation factors (VIF); all values were below 2.0 (range: 1.006–1.054) [25]. Missing data were handled by listwise deletion (outcome item-level missing: 5.0–6.8%; parental attention: 19.0%). No sensitivity analyses were conducted; this is acknowledged as a limitation.

### Ethics statement

This study is a secondary analysis of anonymized, de-identified national data collected in 2013 by KAIMRC in collaboration with NFSP. The original study received institutional ethics review and all participants provided informed consent in accordance with the Declaration of Helsinki. No additional ethics approval was required for this secondary analysis.

## Results

### Sample characteristics

Table 1 presents descriptive statistics for the full sample (N = 10,156) and bullying subsample (N = 4,632). Mean age was 34.44 years (SD = 11.73). Saudi nationals constituted 98.9% of the full sample. Males comprised 52.0% of the full sample and 56.4% of the bullying subsample. Missing data: primary exposure 0%; outcome variables 5.0–6.8%; parental attention 19.0%; all other covariates <3.0%.

### Bullying exposure distribution and outcome prevalence

Approximately 26.1% of the full sample (N = 2,650) reported occasional bullying (’few times’) and 12.8% (N = 1,300) repeated exposure (’many times’). Younger adults (18–31 years) reported the highest frequent bullying rates (49.9%) compared to middle-aged (34.2%) and older adults (15.8%). Males reported higher frequent bullying (36.5%) than females (27.2%). Table 2 presents the full distribution.

**Table 2.** Distribution of bullying frequency and family context variables by gender and age cohort.

| Variable | Male<br>(n=2,611) | Female<br>(n=2,021) | 18–31 yrs | 32–45 yrs | 46–60 yrs |
| --- | --- | --- | --- | --- | --- |
| 'Many times' bullied, n (%) | 833 (36.5%) | 458 (27.2%) | 643 (49.9%) | 441 (34.2%) | 204 (15.8%) |
| 'Few times' bullied, n (%) | 1,428<br>(62.5%) | 1,188<br>(70.6%) | 1,261<br>(48.6%) | 885 (34.1%) | 446 (17.2%) |
| Education <HS: many times (%) | 39.7% | — | — | — | — |
| Education HS: many times (%) | 33.1% | — | — | — | — |
| Education college: many times (%) | 27.8% | — | — | — | — |
| Divorced/separated parents, n (%) | 346 (51.3%) | 328 (48.7%) | 320 (48.0%) | 217 (32.6%) | 129 (19.4%) |
| No parental attention, n (%) | 1,402<br>(61.3%) | 886 (38.7%) | 1,110<br>(48.8%) | 474 (32.8%) | 417 (18.3%) |

*HS, high school.*

Table 3 presents outcome prevalence within the bullying subsample. Sleep disturbance was most prevalent (55.9%), followed by anxiety (26.0%), smoking (44.9%), suicidal ideation (13.0%), and substance use (11.5%).

**Table 3.** Prevalence of adult health outcomes by gender, bullying subsample (N = 4,632).

| Outcome | Male % | Female % | Overall % |
| --- | --- | --- | --- |
| Suicidal ideation | 11.1 | 14.4 | 13.0 |
| Anxiety disorder | 21.6 | 30.4 | 26.0 |
| Sleep disturbance | 50.8 | 62.4 | 55.9 |
| Tobacco smoking | 63.2 | 21.5 | 44.9 |
| Substance use | 6.7 | 15.3 | 11.5 |

### Dose-response associations: logistic regression results

Table 4 presents adjusted odds ratios for all five outcomes. All models were statistically significant (all p < .001). Reference categories: bullying = ’many times’; type = physical; gender = male; age = 18–31 years; parental divorce = no; parental attention = no.

**Table 4.** Binary logistic regression: adjusted odds ratios for adult health outcomes (N = 4,632).

| Predictor | Drug use<br>OR (95% CI) | Smoking<br>OR (95% CI) | Suicidal<br>ideation<br>OR (95% CI) | Sleep<br>OR (95%<br>CI) | Anxiety<br>OR (95%<br>CI) |
| --- | --- | --- | --- | --- | --- |
| Model $\chi^2$ (df) | 186.83*** | 645.84*** | 187.65*** | 77.45*** | 95.82*** |
| Nagelkerke R<sup>2</sup> | 11.8% | 24.7% | 11.8% | 3.3% | 4.5% |
| Classification accuracy | 89.3% | 70.6% | 89.0% | 57.6% | 74.4% |
| **BULLYING FREQUENCY**<br>(ref: many times) |  |  |  |  |  |
| Few times bullied | 0.50 (0.39–0.64)*** | 0.81 (0.67–0.97)* | 0.87 (0.68–1.11) | 0.92 (0.78–1.09) | 0.97 (0.81–1.17) |
| Never bullied | 0.21 (0.05–0.87)* | 0.37 (0.16–0.85)* | 0.45 (0.14–1.45) | 0.39 (0.20–0.76)** | 0.37 (0.16–0.86)* |
| **VERBAL TYPE** (ref: physical) |  |  |  |  |  |
| Religion/belief-targeted | 3.24 (1.92–5.46)*** | 1.33 (0.88–2.01) | 3.01 (1.83–4.97)*** | 0.65 (0.44–0.96)* | 2.39 (1.65–3.46)*** |
| Sexual jokes/gestures | 1.99 (1.04–3.81)* | 1.77 (1.15–2.71)** | 1.99 (1.06–3.72)* | 0.75 (0.50–1.12) | 1.79 (1.16–2.76)** |
| Race/nationality-targeted | 1.50 (0.86–2.60) | 1.21 (0.87–1.68) | 1.83 (1.09–3.09)* | 0.79 (0.58–1.07) | 1.06 (0.74–1.51) |
| **FAMILY CONTEXT** |  |  |  |  |  |
| Divorced/separated parents | 1.19 (0.65–2.19) | 0.65 (0.53–0.79)*** | 0.36 (0.22–0.58)*** | 0.72 (0.59–0.89)** | 0.70 (0.56–0.88)** |
| Parental attention (yes) | 0.45 (0.30–0.67)*** | 0.99 (0.84–1.18) | 2.18 (1.56–3.04)*** | 0.96 (0.81–1.14) | 1.00 (0.84–1.19) |
| **DEMOGRAPHICS** |  |  |  |  |  |
| Female (ref: male) | 2.71 (2.26–3.31)*** | 6.55 (5.81–7.32)*** | 0.69 (0.54–0.88)** | 0.63 (0.54–0.73)*** | 0.63 (0.54–0.74)*** |
| Age 32–45 years | 0.76 (0.55–1.05) | 0.78 (0.62–0.97)* | 1.85 (1.30–2.65)** | 1.19 (0.97–1.46) | 0.69 (0.55–0.87)** |
| Age 46–60 years | 0.97 (0.67–1.41) | 0.91 (0.72–1.14) | 1.22 (0.81–1.82) | 1.13 (0.91–1.40) | 0.92 (0.72–1.17) |

All ORs adjusted. \* $p < .05$ ; \*\* $p < .01$ ; \*\*\* $p < .001$ . Per-model N varies due to listwise deletion on parental attention variable (19.0% missing).

#### Dose-response gradient (H1 and H2)

H1 and H2 were confirmed across behavioral outcomes. For substance use, respondents bullied ’few times’ had 50% lower odds than those bullied ’many times’ (OR = 0.50, 95% CI 0.39–0.64, p < .001), and those never bullied had 79% lower odds (OR = 0.21, 95% CI 0.05–0.87, p = .036). For tobacco smoking, ’few times’ (OR = 0.81, p = .021) and ’never bullied’ (OR = 0.37, p = .015) showed significantly lower odds than the ’many times’ reference, confirming the dose-response gradient.

#### Religion-targeted victimization

Religion-targeted verbal bullying was the strongest predictor of suicidal ideation (OR = 3.01, 95% CI 1.83–4.97, p < .001), substance use (OR = 3.24, 95% CI 1.92–5.46, p < .001), and anxiety disorder (OR = 2.39, 95% CI 1.65–3.46, p < .001), independently of bullying frequency.

#### Parental supervision and family context

Parental attention was a significant protective factor for substance use (OR = 0.45, 95% CI 0.30–0.67, p < .001) but positively associated with suicidal ideation (OR = 2.18, 95% CI 1.56–3.04, p < .001). Living with divorced or separated parents was associated with lower odds of suicidal ideation (OR = 0.36), smoking (OR = 0.65), sleep disturbance (OR = 0.72), and anxiety (OR = 0.70).

### Other analyses

Interaction models provided partial support for H3: bullying-health associations were significantly stronger among socioeconomically disadvantaged respondents (interaction term p < .05 across behavioral outcomes). Table 5 confirms no multicollinearity (all VIF < 2.0).

**Table 5.**
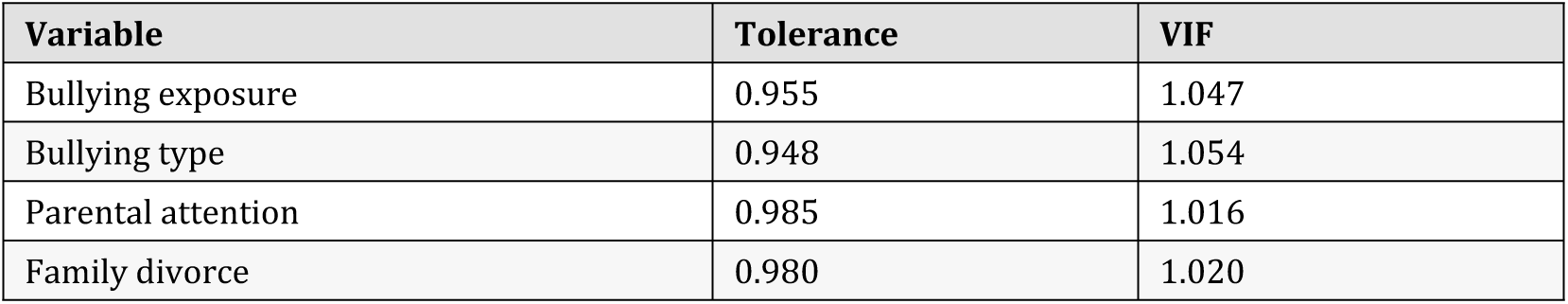
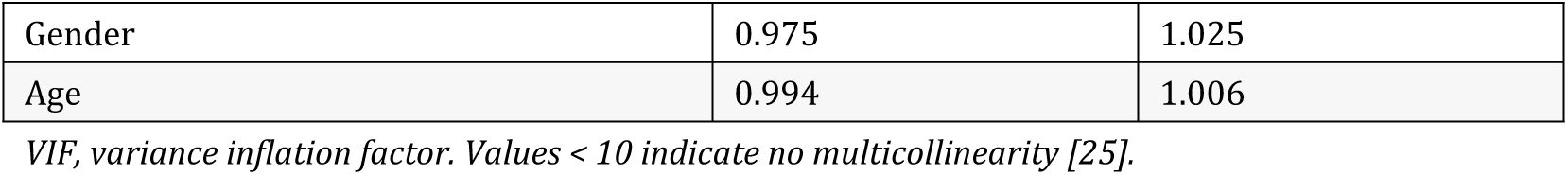
Collinearity statistics.

| Variable | Tolerance | VIF |
| --- | --- | --- |
| Bullying exposure | 0.955 | 1.047 |
| Bullying type | 0.948 | 1.054 |
| Parental attention | 0.985 | 1.016 |
| Family divorce | 0.980 | 1.020 |
| Gender | 0.975 | 1.025 |
| Age | 0.994 | 1.006 |

VIF, variance inflation factor. Values < 10 indicate no multicollinearity [25].

## Discussion

### Key findings

This study provides national dose-response evidence supporting all three hypotheses. H1 was confirmed: any bullying exposure was associated with higher odds of adverse adult health outcomes. H2 was confirmed across behavioral outcomes: increasing bullying frequency was associated with progressively higher odds of smoking and substance use. H3 was partially confirmed: socioeconomic disadvantage amplified associations, while parental supervision was protective against substance use.

### Dose-response gradient and cumulative health risk

The dose-response pattern — most pronounced for tobacco smoking (Nagelkerke R² = .247) and substance use (OR = 2.71, 95% CI 2.26–3.31) — establishes that peer victimization operates as a continuously graded health risk rather than a binary threshold. This is consistent with models in which chronic social adversity dysregulates HPA axis function, contributes to allostatic load, and elevates vulnerability to behavioral coping adaptations [10,26]. These findings extend recent longitudinal evidence from Takizawa et al. [5], Blanchflower and Bryson [7], and Momose and Ishida [27], contributing cross-national evidence from a non-Western context.

### Religion-targeted victimization as an independent risk factor

Religion-targeted verbal bullying was the strongest independent predictor of suicidal ideation (OR = 3.01), substance use (OR = 3.24), and anxiety (OR = 2.39). In Saudi Arabia, where religious identity is embedded in social legitimacy, victimization targeting religious practice may represent identity-based stigmatization with amplified psychological consequences [28,29]. Prevention programs addressing only physical or frequency-based bullying without attention to identity-targeted forms may substantially underestimate the mental health burden generated [3].

### Structural amplification: socioeconomic disadvantage and health inequality

The amplification of bullying-health associations among socioeconomically disadvantaged respondents is consistent with fundamental cause theory [16,17]: disadvantage both increases exposure risk and diminishes protective resources, producing compounded inequalities that require equity-focused policy responses beyond school-level programs [17,18].

### Parental supervision: protective and paradoxical effects

Parental supervision was significantly protective against substance use (OR = 0.45, p < .001). The positive association with suicidal ideation (OR = 2.18) is most parsimoniously explained by reactive parenting dynamics in a cross-sectional design: parents who increased monitoring in response to a child’s visible distress would produce precisely this positive association [30]. The 19.0% missing data rate warrants treating these findings as hypothesis-generating.

### Gender-differentiated health pathways

The adjusted OR for female smoking (OR = 6.55) reflects a statistical suppression effect: after adjustment, bullied women’s conditional smoking risk substantially exceeds prediction given strong social sanctions against female smoking in Saudi Arabia [31]. Bivariate female smoking prevalence (21.5% vs. 63.2% male) confirms this reflects excess conditional risk, not high absolute rates. Lower female adjusted odds for suicidal ideation (OR = 0.69) and anxiety (OR = 0.63) despite higher bivariate prevalence likely reflect differential diagnostic ascertainment and help-seeking norms [32].

### Public health implications

These findings support repositioning childhood bullying as a population health priority requiring multi-level prevention:

- School-based programs should adopt frequency- and content-sensitive approaches with specific protocols for identity-targeted victimization.
- Family support services emphasizing emotional warmth alongside behavioral supervision represent a modifiable protective pathway [30,33].
- Prevention resources should be preferentially allocated to socioeconomically disadvantaged communities [17,18].
- Bullying prevention should be integrated into Saudi Arabia’s Vision 2030 national youth health agenda aligned with WHO ACE surveillance [2,34].

### Limitations

Several limitations apply. First, the retrospective cross-sectional design precludes causal inference and introduces recall bias risk (positive bias direction). Second, the ACE-IQ parental attention item captures behavioral monitoring rather than emotional warmth. Third, the parental attention variable had 19.0% missing data (possible attenuation bias). Fourth, co-occurring ACEs were not fully adjusted for (residual confounding). Fifth, no formal design-effect corrections for cluster sampling were applied. Sixth, all outcomes were self-reported. Seventh, no sensitivity analyses were conducted.

### Interpretation

The consistency of behavioral health dose-response patterns, independence of religion-targeted victimization effects, and structural moderation by socioeconomic disadvantage constitute a coherent pattern that is difficult to explain by recall bias or confounding alone, though causal inference requires longitudinal replication. Findings are consistent with and extend the existing bullying-health literature [5,7,27] to a non-Western, Muslim-majority context.

### Generalisability

The nationally representative Saudi sampling frame supports generalisability to the Saudi adult population aged 18–60 years. Generalisation to other GCC or Muslim-majority contexts should be considered cautiously given differences in social structures, gender norms, and healthcare utilization patterns.

## Conclusions

This study establishes childhood bullying as a cumulative, graded public health risk in Saudi Arabia, with increasing victimization frequency associated with progressively higher odds of adverse adult mental and behavioral health outcomes. The dose-response gradient — strongest for substance use (OR = 2.71) and tobacco smoking (R² = .247) — provides epidemiological evidence that peer victimization should be conceptualized as a frequency-dependent health exposure within ACE frameworks. Religion-targeted verbal bullying is a distinct and potent independent risk factor. Socioeconomic disadvantage amplifies associations, positioning bullying as a mechanism of structural health inequality requiring equity-focused policy. Findings contribute population-level ACE evidence from the underrepresented Gulf region, support integration of bullying prevention into Saudi Arabia’s Vision 2030 health agenda, and validate the WHO ACE-IQ as a productive cross-national surveillance tool. Future research should employ longitudinal cohort designs, validated parental warmth instruments, and extend outcomes to physical health endpoints.

## Data Availability

The data underlying this study are not publicly available due to institutional data governance and participant confidentiality agreements governing national health surveillance data in the Kingdom of Saudi Arabia. The ACE-IQ dataset used in this study was collected by the King Abdullah International Medical Research Center (KAIMRC) in collaboration with the National Family Safety Program (NFSP), under the Health Affairs of the Ministry of National Guard. As a national health surveillance dataset containing sensitive data on adverse childhood experiences, the data are subject to formal institutional access controls. Researchers wishing to access the data may submit a formal data access request directly to KAIMRC at www.kaimrc.med.sa. This constitutes a documented legal and ethical restriction consistent with the journal's data availability exemption policy.

## Acknowledgments

The author acknowledges the King Abdullah International Medical Research Center (KAIMRC) and the National Family Safety Program (NFSP), Ministry of National Guard Health Affairs, Saudi Arabia, for providing access to the ACE-IQ national dataset as part of the doctoral research program at South Dakota State University.

## Supporting information

**S1 Checklist. STROBE checklist for cross-sectional studies.** Completed checklist confirming all 22 STROBE items are addressed in this manuscript.

**S1 Fig. Study participant flow diagram.** High-resolution JPEG file (300 DPI) of Fig 1 as a separate upload file named Fig1.jpg.

## Notes

### Competing Interest Statement

The authors have declared no competing interest.

### Author Declarations

This study is a secondary analysis of fully anonymized and de-identified national data originally collected in 2013 by the King Abdullah International Medical Research Center (KAIMRC) in collaboration with the National Family Safety Program (NFSP), under the Health Affairs of the Ministry of National Guard, Saudi Arabia. The original data collection was conducted under institutional ethics review and approval granted by KAIMRC and through the NFSP official research application and approval process. All participants in the original study provided written informed consent prior to participation in accordance with the Declaration of Helsinki. The secondary analyst (Dr. Dalal A. Bin Hamdan, Ph.D.) accessed only fully de-identified data with no individual-level identifiers. As this study constitutes a secondary analysis involving no direct interaction with human participants and using exclusively pre-existing, fully de-identified national surveillance data, no additional IRB or ethics committee approval was required for this secondary analysis. This exemption is consistent with standard international research ethics guidelines for secondary analysis of anonymized datasets.

